# A modified Delphi study to determine healthcare consumer priorities and consensus for the use of genomics and precision medicine approaches in pregnancy healthcare and research

**DOI:** 10.64898/2026.09.29.26363624

**Authors:** Tegan Grace, Samantha Hoskins, Kirsty Pringle, Melinda Cruz, Keren Ludski, Leila Usher, Nafiseh Ghafournia, Craig E. Pennell

**Affiliations:** School of Medicine and Public Health, College of Health, Medicine and Wellbeing, The University of Newcastle; Reproductive and Family Health Research Program, Hunter Medical Research Institute; Mother and Babies Research Centre, University of Newcastle; School of Biomedical Sciences and Pharmacy, College of Health, Medicine and Wellbeing, The University of Newcastle; NHMRC Clinical Trials Centre, Faculty of Medicine and Health, University of Sydney; NICU Lived Network; Red Nose; School of Humanities, Creative Industries and Social Sciences, University of Newcastle

**Author notes:** **Corresponding Author** Dr Tegan Grace, RFHRP Level 3 East, Hunter Medical Research Institute, Lot 1 Kookaburra Avenue, New Lambton Heights, New South Wales, Australia, 2305 E.

**Keywords:** Consumer engagement, Precision medicine, Delphi study

## Abstract

**Background:** Pregnancy is a uniquely complex ethical and clinical context in which precision medicine approaches, including genomics-driven risk prediction tools, biomarker-based stratification, and data-intensive analytics, are rapidly evolving. There is a need for community consultation on the ethical, social, and practical implications of precision medicine in pregnancy research and healthcare.

**Aim:** The primary aim of this study was to engage healthcare consumers to build consensus on the acceptability of precision medicine approaches in pregnancy research and healthcare. We also sought to identify consumer priorities for future research.

**Methods:** A two-round modified Delphi study was conducted to determine consumer opinions on the use of precision medicine approaches in pregnancy research and healthcare. Seven in-person workshops (round one) were held, with purposeful recruitment to ensure a representative group of healthcare consumers who engage with the healthcare system during pregnancy. These included first-time parents, culturally and linguistically diverse women, Aboriginal and Torres Strait Islander women, and women and their partners who had experienced previous pregnancy complications. Recruitment was conducted through established advocacy networks, and each workshop was facilitated by representatives from each organization. A second-round online survey, based on workshop content, was completed to assess the acceptability of precision medicine and to determine consumer priorities for future precision medicine research on pregnancy health.

**Results:** Precision medicine research and the implementation of new technologies in clinical care were acceptable to participants; however, the need for transparency and trust between participants and researchers regarding the use and storage of genetic material was highlighted, particularly among Aboriginal and culturally and linguistically diverse participants. Outcomes related to survival and perinatal complications (stillbirth, preterm birth, and labor complications) were the highest-rated consumer research priorities, along with maternal mental health and breastfeeding. When asked whether participants would like to see PM available as part of routine pregnancy care for conditions with available treatments, complete consensus (100%) was reached.

**Conclusion:** Healthcare consumers strongly support the use of precision medicine in pregnancy research and its translation into routine clinical care, particularly when effective interventions are available. However, implementation must be underpinned by transparency, trust, culturally appropriate governance, and clear communication about the collection, storage, and use of genetic material. These findings provide a consumer-informed framework for prioritizing future precision medicine research and guiding its responsible translation into pregnancy care.

**Key Points:** Australian healthcare consumers were broadly supportive of precision medicine approaches to pregnancy research and healthcare

Implementation of precision medicine into research and healthcare must be underpinned by robust and transparent governance, clear communication and culturally appropriate guidance.

## Introduction

Precision medicine (PM) approaches, including genomics-driven risk prediction, biomarker-based stratification, and data-intensive analytics, are increasingly proposed as solutions to persistent challenges in obstetrics (1-6). However, pregnancy presents a unique and complex ethical context in which emerging technologies are developed and applied, with implications not only for the pregnant person but also for the fetus and the family (2, 3, 7). This is particularly important when risk stratification screening may result in prognostic uncertainty and is conducted under time-sensitive conditions (3). Experience with existing technologies and the rapid expansion of prenatal genomic testing underscore the importance of consumer engagement to ensure that emerging technologies are introduced with adequate professional understanding of patient needs, to avoid generating anxiety, to uphold informed consent, and to ensure equitable access (2, 3). While PM research in antenatal care continues to grow, it is recognized that technological advances and clinical promise alone are insufficient to justify implementation without meaningful, concurrent consumer engagement to assess acceptability, perceived benefit, risk, and trust (1, 2).

Meaningful consumer engagement early in research is essential for guiding the development and translation of PM tools and for ensuring ethical, patient-centered implementation practices. Consumer engagement has been shown to improve the relevance of research aims, ensure that outcomes are meaningful to patients, and guide the development of acceptable communication strategies for risk, data governance, and consent (8). This is particularly important in research involving genomics and the future applications of new technologies in pregnancy care (1). There is a clear need for community consultation on the ethical, social, and practical implications of genomics and precision medicine in pregnancy healthcare (1, 9, 10).

Current Australian policy emphasizes meaningful engagement with healthcare consumers throughout the design, conduct, and translation stages of research (8). The objectives of this study were therefore to (a) engage Australian healthcare consumers to develop consensus on the acceptability of genomics-enabled precision medicine approaches in research and clinical care, and (b) identify consumer-defined priorities, values, and concerns to inform the design, governance, and future priorities of precision medicine research in maternal-fetal medicine.

## Methods

### Study Design

This study involved three years of consultation with two of the largest consumer-led not-for-profit organizations in Australia supporting families affected by pregnancy complications, including preterm birth (Miracle Babies) (11) and sudden unexpected death during pregnancy and infancy (Red Nose) (12). Ongoing discussions with key healthcare consumer representatives from these organizations highlighted the need for precision medicine research to inform pregnancy healthcare practice. As precision medicine approaches to pregnancy research and healthcare are likely to impact a broader range of antenatal health outcomes, recruitment was expanded to include a larger, more representative group of consumers. A modified Delphi method was selected for this study because it allowed thematic analysis of workshop discussions and quantitative analyses of online surveys to determine consensus. The Delphi approach has been extensively used to anticipate issues in areas of developing knowledge, technology, or interventions, thereby facilitating appropriate governance and priority setting (1). This formal technique is used to obtain consensus within a group considered to have expertise in a specific area (9), using a mixed-methods approach that combines qualitative and quantitative methods. Through a series of ‘rounds,’ the group is asked for their views on an issue or topic about which they have experience or expertise. These answers then form the basis of the next ‘round,’ where participants are given the opportunity to reflect on their answers and the answers of others before providing further input. The overarching aim is to reach consensus on the topic or issue.

The modified Delphi method involved two rounds: an in-person workshop (round one) and an online survey (round two). Seven workshops were held using purposive recruitment to ensure a diverse group representative of healthcare consumers who engaged with the healthcare system during pregnancy, including first-time parents, culturally and linguistically diverse women, Aboriginal and Torres Strait Islander women, and women and their partners who had experienced previous pregnancy complications. In line with other Delphi studies, the recruitment focus was on participant diversity to ensure good representation, rather than on a sample size to achieve statistical significance (13). Workshops included a short presentation on precision medicine, followed by questions to explore healthcare consumers’ views on its use in pregnancy research and healthcare. Following established guidelines (14, 15), the workshops used open-ended questions to generate ideas and allow free responses. Questions about the use of precision medicine approaches in pregnancy healthcare, particularly genomics in pregnancy, were addressed through group discussion, with answers recorded by scribes.

Questions asked whether participants agreed with research using maternal and paternal DNA and/or fetal cell-free DNA (cfDNA) extracted from maternal blood samples to develop precision medicine screening tools for aneuploidy, preterm birth, fetal growth restriction, or other pregnancy issues. Participants were invited to list as many ‘pros’ and ‘cons’ of this type of testing as they liked. Participants were also asked to identify the most important pregnancy-related health issues or conditions for future research focus. The second-round survey was sent via the Research Data Capture database (REDCap) and focused on the areas of pregnancy health that participants believed were most important for future research, based on lists compiled from the workshops.

### Participants

#### Recruitment

Participants were recruited from the John Hunter Hospital in Newcastle, NSW; Miracle Babies; Red Nose; the Awabakal Ltd Mums and Bubs Program (16); and the Gomeroi Gaaynggal study (17). A research midwife at the John Hunter Hospital antenatal clinic identified first-time parents and provided information sheets about the workshops. The Multicultural Health Liaison Officer (MHLO) within the hospital’s Mothers, Obstetrics and Multicultural Support (MOMS) program identified culturally and linguistically diverse (CALD) women interested in participating. The Hunter New England Health Care Interpreter Service provided translated information and consent forms, as well as in-person interpreters. The MHLO facilitated the workshops for CALD women.

Mothers and fathers with a history of pregnancy complications or loss were identified through the networks of the organizations Red Nose and Miracle Babies. Each workshop was facilitated by an organization representative. The Gomeroi Gaaynggal Study (17) Steering Committee, including Elders and participants, was approached by the Study Director, who has established networks within the Tamworth community. The workshop was facilitated by an Aboriginal research officer and the Study Director. Aboriginal women from the HNELHD were approached by an Aboriginal Obstetrician who worked with the local Aboriginal Controlled Community Health Organization’s maternal health program and also facilitated the workshop.

In line with the Healthcare Consumer NSW guidelines, participants received a voucher equivalent to $42/hour for attending the half-day workshop and completing the online survey (18).

### Data Analysis

Scribed data from the workshops were grouped into positive, negative, and contingent responses and then thematically analyzed in NVivo 12 (QSR International). Pregnancy-related health issues of concern were included in the second-round survey if they were mentioned by at least four of the seven groups. The survey results were used to determine healthcare consumers’ rankings of the top five research priorities for pregnancy research. Rankings were converted to weighted scores (1st□=□5 points, 5th□=□1 point) to generate an overall priority ranking.

## Results

Seventy-six people participated in the workshops, including 19 (25%) males (M = 37.1 years, SD = 3.1 years) and 57 (75%) females (M = 36.00 years, SD = 6.69 years). Participants included nulliparous women and their partners (n = 12); women with a history of pregnancy complications resulting in stillbirth or sudden unexpected death of an infant (SUDI) (n = 13); women with a history of pregnancy complications resulting in preterm birth (n = 9); fathers with various pregnancy histories (n = 13); Aboriginal women from the Gomeroi Gaaynggal cohort (17) in Tamworth (n = 5); Aboriginal women from the Hunter New England Local Health District (n = 14); and culturally and linguistically diverse (CALD) women (n = 10).

### Acceptability of precision medicine in pregnancy research

Workshops began with a brief presentation on PM. Participants were encouraged to ask questions to clarify their understanding. The workshop discussion focused on participants’ views on the use of maternal and paternal DNA and fetal cell-free DNA (cfDNA) in research, and on integrating PM into routine pregnancy healthcare. The use of maternal, paternal, and fetal cfDNA for research was not seen as prohibitive; however, the need for transparency and trust between participants and researchers regarding the use and storage of genetic material was highlighted, particularly among Aboriginal and CALD participants. While the need for transparency and the historical foundation for distrust among some groups were discussed, consumers in these groups also expressed that research on new technology should be inclusive of minority groups to ensure they are equally involved in the design, implementation, governance, and consent processes. Transparency and communication were key.

*“A strong focus on open communication and building long-term relationships is a must within the Aboriginal community. It’s all about the yarn and how we engage with those providing healthcare” (F, age 30-35)*.

The potential to use fetal cfDNA for precision medicine research was acceptable, with participants stating it was ‘less invasive’ to obtain fetal DNA from maternal blood rather than from fetal sources, ‘as long as it was safe to do so’. Paternal DNA was discussed as a potential limiting factor in precision medicine research if the father was unavailable, unwilling to participate in research, unwilling to give blood, or had reservations about the use of their DNA for legal reasons related to proof of paternity, substance use, or other concerns. Other options for DNA collection, such as saliva, were discussed as viable alternatives if participants were unwilling to provide blood samples.

### Potential benefits of precision medicine in routine pregnancy healthcare

The use of precision medicine approaches in routine pregnancy care was discussed, with participants invited to share any thoughts, positive or negative, about PM tools becoming available as part of regular pregnancy healthcare. Qualitative analyses revealed three main benefits of this approach: ‘knowledge is power’, ‘time to prepare’, and ‘earlier intervention’. Participants viewed PM approaches to pregnancy healthcare as a positive step if they allowed parents to be better prepared for potential adverse outcomes during pregnancy. Participants noted that ‘knowledge means you can better prepare and get your head around it’, and that this may also result in less stress for parents as they have an ‘exact picture of what’s going on.’ In addition to mental preparedness, the ability to plan and prepare in a practical sense was viewed as important.

*‘… even if you can’t do anything about it you can better prepare yourself’ (M, age 25-30)*.

This included arranging additional support, moving closer to healthcare facilities if living in a remote or regional area, scheduling bed rest or additional levels of care, and gaining a better understanding of the newborn’s postnatal needs. The possibility of early intervention to treat or reduce the risk of pregnancy-related issues, such as preterm birth (where treatments are available), was also seen as a potential benefit. This was particularly evident when participants thought that PM approaches could better target those more likely to respond to intervention.

### Potential limitations of precision medicine in routine pregnancy healthcare

Participants noted several potential drawbacks to PM, including ‘increased stress’, ‘guilt’, and ‘where do you stop?’. Increased stress on parents, particularly expectant mothers, when screening or test results showed a higher risk for complications was a common theme in responses. This was identified as the largest potential negative of screening tools for pregnancy complications, although participants also noted that multiple screening tools are already used in pregnancy healthcare and that the potential for precision medicine to make them more accurate was appealing. Participants mentioned the possibility that parents might feel guilty if genomic testing revealed they have ‘faulty genes’ that increase the risk of pregnancy complications. The question of ‘where do you stop’ was also discussed, with participants expressing concern about how parents might interpret results from new screening technologies. One potential issue was whether an indication of higher risk might lead to a higher rate of pregnancy termination if some parents were not willing to go through with a complicated pregnancy or did not want the potential postnatal complications.

### Incorporating precision medicine into routine pregnancy healthcare

When asked about the implications of incorporating PM into routine healthcare, participants indicated that certain stipulations should be met. ‘Choice’, ‘equality of healthcare’, and ‘follow-up support’ for parents following any screening or diagnosis were highlighted. Participants noted that if precision medicine were to become a standard of treatment, it should be an option for parents, not mandatory.

*“I think that if it was routine to ask if this is something you can opt into as part of routine pregnancy care, then that would be great. But I would not like it to be imposed*.*” (F, age 25-30)*

Participants were also concerned that the cost of such technology would be prohibitive for some parents, noting that it was important not to create a two-tiered health system in which parents who could not afford testing were unable to access available healthcare advancements and technologies due to out-of-pocket costs. Support and resources for parents when test results indicate an increased risk of complications were seen as an important part of implementing any new technology or screening tools.

*During all my pregnancies, we used the private healthcare system and were offered multiple tests. It wouldn’t matter to me personally whether they were $ 1,000 or $ 2,000, but this could cause financial stress for other families. (M, age 30-35)*

Consumers also wanted healthcare providers to ‘explain clearly, answer questions, and provide some sort of support network’ when results were given, and participants emphasized the need for a care plan and follow-up for the patient, including mental health or social support where needed. The concept of continuity of care was also highlighted, as participants noted that it would be better if the patient knew and trusted their caregiver when deciding whether to pursue additional screening or tests.

### Priorities research areas for pregnancy research

From the seven workshops, a list of 52 pregnancy-related health issues was compiled. Pregnancy health issues mentioned in the workshops by at least four of the seven groups, constituting a ‘majority’ agreement (1), were included in the online survey, which presented a list of 10 issues. Participants were asked to rank their top five priorities. Rankings were converted to weighted scores (1st□=□5 points to 5th□=□1 point) to generate an overall priority ranking (Figure 1). A total of 51 online surveys (68%) were completed. Outcomes related to survival and perinatal complications (stillbirth, preterm birth, and labor complications) emerged as the highest consumer priorities, with maternal mental health also receiving a high weighted rank. Breastfeeding support was rated the fifth-highest priority, while more specific health conditions, including hypertensive disorders, gestational diabetes mellitus, fetal growth, and existing maternal comorbidities, received similar weighted scores. These findings highlight areas where precision medicine approaches are most likely to align with consumer expectations and perceived clinical value.

**Figure 1.**
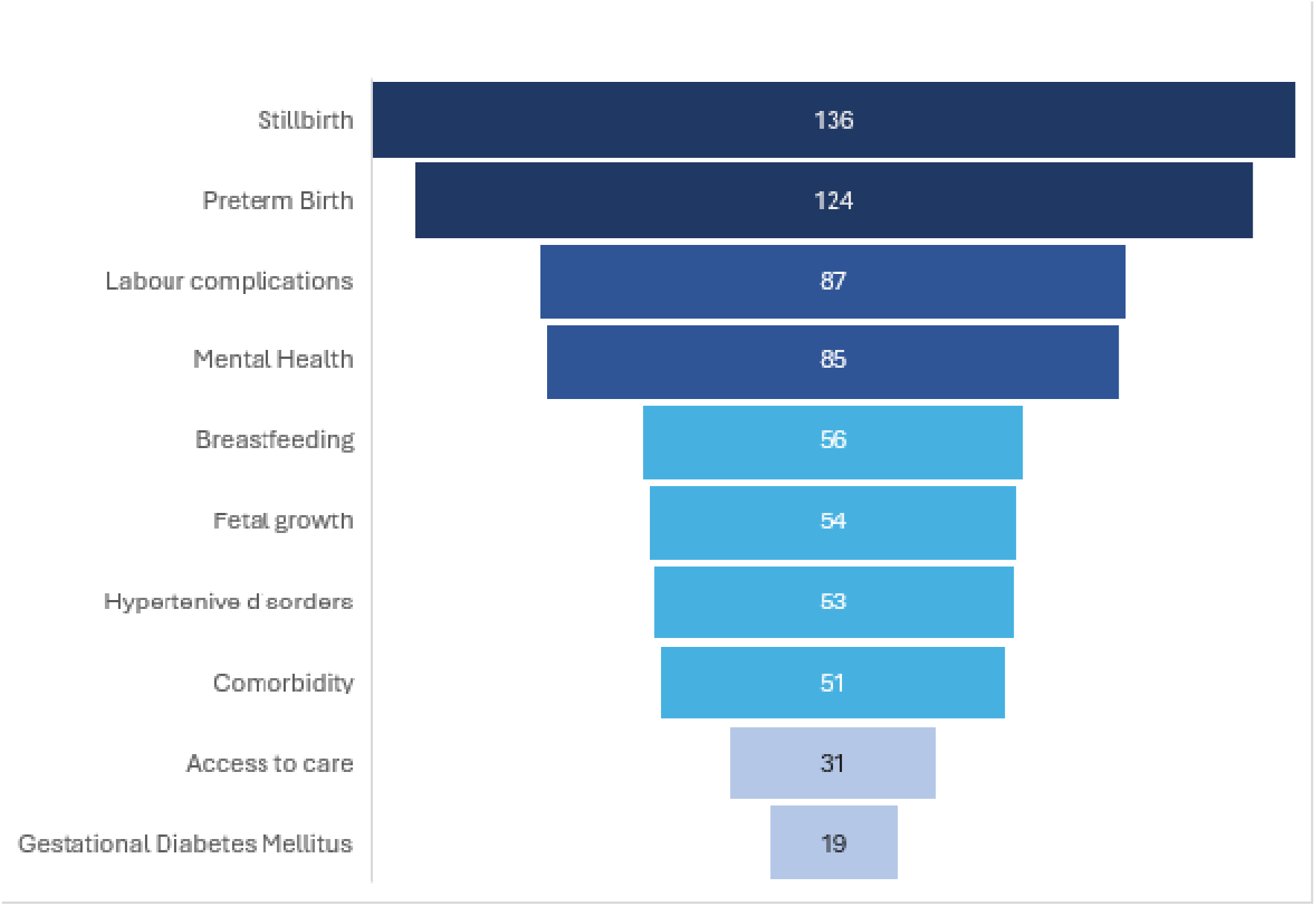
Consumer identified priorities for precision medicine research in pregnancy: weighted scores

The top-ranked pregnancy health issue was stillbirth, with a total weighted score of 136 points. Preterm birth ranked second with a weighted score of 124 points. Labor complications and maternal mental health were weighted at 87 and 85 points, respectively. Breastfeeding (56 points) was the fifth-highest ranked issue, with fetal growth (54 points), hypertensive disorders (53 points), and maternal comorbidities (51 points) ranked within a similar bracket. Access to care (31 points) and gestational diabetes mellitus (19 points) were ranked as the ninth- and tenth-most important health concerns by consumers. A final question in the online survey asked participants whether they would like to see PM available as part of routine pregnancy care for conditions where treatments are available, and complete consensus (100%) was reached.

## Discussion

### Precision medicine in pregnancy research

Our findings indicate that, when researchers were clear and upfront about the use and storage of genetic material, consumers were supportive of PM in pregnancy research. Confidentiality and participant autonomy were especially important in research involving vulnerable populations, particularly Aboriginal and culturally and linguistically diverse consumers. These findings align with a recent systematic review that identified privacy and security as the main concerns for patients regarding ethical issues in PM (10).

### Precision Medicine in routine pregnancy care

Consumers in our study expressed the potential benefits of PM in routine pregnancy care, including increased knowledge (‘knowledge is power’), preparation (‘time to prepare’), and potential intervention (‘earlier intervention’). Discussion of these benefits included increased parents’ ability to make informed decisions about their care, including any available interventions; organizing work and other family commitments where necessary; and the benefit of earlier detection of complications or risk. Earlier detection of risk and subsequent intervention were perceived as beneficial for health outcomes and overall healthcare costs for both families and the healthcare system (6). While the benefits were seen to generally outweigh the potential drawbacks, consumers also highlighted the need for the healthcare system to support the integration of new technologies in a fair and equitable manner. Consumers were aware of the potential for inequity and expressed that new technologies should be made accessible to all families, not only those who could afford out-of-pocket expenses. These responses were in line with recent Australian research that reported over 60% of parents who were not utilizing currently available genomic testing services rated cost as an important factor in their decision (19), and with a recent systematic review that listed economic impact as the second most mentioned ethical concern of PM in pregnancy care (10).

The potential for new screening and technology to increase parental stress and guilt regarding diagnoses or risk was discussed in the workshops, with consumers expressing concern about negative or high-risk results. These concerns are similar to those reported for PM in other populations (10). Consumers highlighted the need for healthcare providers to ensure support, both initially and on an ongoing basis, for detected risks or complications. In the current landscape of pregnancy care in Australia, this raises important issues for implementing new screening tools. Consistency in the application of new technology, the development of national professional guidelines, professional training, and standardization may be challenging if testing and results are conducted by commercial companies. The implementation of new technologies and screening tests should be undertaken with ongoing consumer engagement, particularly among vulnerable populations.

### Priorities for future research focus

The second round of the Delphi study identified the highest priorities for future research. Consumers identified stillbirth, preterm birth, labor complications, maternal mental health, and breastfeeding as the top five priority areas for research. The benefits of precision medicine approaches for improving the detection, intervention, and treatment of these conditions should be a focus of future consumer-driven precision medicine research. Conventional approaches to these conditions have relied heavily on population-based risk stratification, single-disciplinary approaches, and standardized clinical surveillance (5, 20). However, these approaches may not account for the marked biological and contextual heterogeneity across pregnancies or effectively elucidate meaningful variation in disease susceptibility, progression, and treatment response, particularly where objectivity and observer bias can influence outcomes (5, 21). Precision medicine approaches to identifying women at high risk of specific conditions involve integrating data across multiple domains – including genomics, molecular biomarkers, clinical parameters, imaging, and environmental exposures – through continually evolving data-driven platforms, including machine learning (6, 22). Consumers emphasized preventing severe outcomes, including mental health alongside physical outcomes, and ensuring that emerging precision medicine approaches improve real-world pregnancy care. Through improved discrimination and individualized treatment, precision medicine approaches have the potential to improve these consumer-identified health outcomes.

### Strengths

Although we recognize that not all family dynamics were represented in the groups, we used a purposeful recruitment strategy to include healthcare consumers from multicultural backgrounds and with diverse pregnancy experiences. We acknowledge that there were small numbers of LGBTIQ+ consumers in the study, and families that rely on egg or sperm donation may hold differing views on research and healthcare, such as precision medicine, which relies heavily on genetics for risk screening and stratification.

### Limitations

As outlined by Rowe and Wright (23), one of the four criteria for strict adherence to the Delphi technique is anonymity, which allows participants to respond without undue social pressure. Although the first round of this modified Delphi study was conducted in a workshop and therefore did not allow for complete anonymity, the researchers carefully worded questions to reassure participants that there were no ‘right or wrong answers’ and used open-ended questions to allow freedom of response. Another limitation is that a consensus study may underreport minority groups’ views on important issues if specific concerns are voiced by fewer people. We sought to address this through purposeful recruitment of Aboriginal and CALD families.

## Conclusion

Healthcare consumers strongly support the use of precision medicine in pregnancy research and its translation into routine clinical care, especially when effective interventions are available. However, implementation must be grounded in transparency, trust, culturally appropriate governance, and clear communication about the collection, storage, and use of genetic material. These findings offer a consumer-informed framework for prioritizing future precision medicine research and guiding its responsible translation into pregnancy care.

## Data Availability

Data from this study is available upon request to the corresponding author

## Data Availability

Data from this study is available upon request to the corresponding author

## Ethics Approval

This study was approved by the Hunter New England Local Health District Human Research Ethics Committee (2020/ETH01175).

## Patient Consent

All participants of this study were consented via informed written consent. All participants were advised they could withdraw from the study at any time.

## Author Contributions

TG, KL, MC, CP, LU, NG, KP contributed to the study design and were directly involved in facilitating the workshops, SH attended as a consumer. TG analysed the qualitative and quantitative data. TG, CP, KL, MC, LU, NG, SH and KP contributed to the final manuscript.

## Acknowledgements

We pay our respects to the Awabakal and Kamilaroi people, the traditional custodians of the lands on which our workshops were held, and we gratefully acknowledge the Gomeroi Gaaynggal Advisory Committee, Professor Kirsty Pringle, the Gomeroi Gyaanggal Study Director and Rachel Hatfield, for facilitating the Tamworth workshop. We thank Dr Leila Usher from Awabakal for facilitating the Aboriginal women’s workshop and father’s workshop in Newcastle, and Nafiseh Ghafournia from the MOMs Program for her assistance in recruitment and facilitation of the CALD workshop. We acknowledge and are grateful for the work of facilitators Kerin Ludski (Red Nose) and Melinda Cruz (Miracle Babies) in recruiting participants from their networks and facilitation of workshops, and Oyepeju Onifade, Saije Morosin and Alyssa Lochrin for scribing and assisting in the running of workshops. We are extremely grateful to the participants of this study for their time and dedication to improving the health and wellbeing of future generations.

